# Conceptualizing centers of excellence: A global evidence

**DOI:** 10.1101/2021.03.17.21253854

**Authors:** Tsegahun Manyazewal, Yimtubezinash Woldeamanuel, Claire Oppenheim, Asrat Hailu, Mirutse Giday, Girmay Medhin, Anteneh Belete, Getnet Yimer, Asha Collins, Eyasu Makonnen, Abebaw Fekadu

## Abstract

**Objective:** Globally, interest in excellence has grown exponentially, with public and private institutions shifting their attention from meet targets to achieving excellence. Centers of Excellence (CoEs) are standing at the forefront of healthcare, research, and innovations responding to the world’s most complex problems. However, their potential is hindered by conceptual ambiguity. We conducted a global synthesis of the evidence to conceptualize CoEs.

**Design:** Scoping review, following Arksey and O’Malley’s framework and methodological enhancement by Levac *et al* to analyze the evidence and the PRISMA-ScR to guide the retrieval and inclusion of the evidence.

**Data sources:** PubMed, Scopus, CINAHL, Google Scholar, and the Google engine from their inception to 01 January 2021.

**Eligibility:** Papers that describe CoE as the main theme, which could be defining, theorizing, implementing, or evaluating a CoE.

**Results:** The search resulted in 52,161 potential publications, with 78 articles met the eligibility criteria. The 78 articles were from 33 countries, of which 35 were from the United States of America, 3 each from Nigeria, South Africa, Spain, and India, and 2 each from Ethiopia, Canada, Russia, Colombia, Sweden, Greece, and Peru. The rest 17 were from various countries. The articles involved six thematic areas - healthcare, education, research, industry, information technology, and general concepts on CoE. The analysis documented success stories of using the brand “Center of Excellence” - an influential brand to stimulate technical skills, innovation, and technology. We identified 12 essential foundations of CoE - specialized expertise; infrastructure; innovation; high-impact research; quality service; accreditation or standards; leadership; organizational structure; strategy; collaboration and partnership; sustainable funding or financial mechanisms; and entrepreneurship.

**Conclusions:** CoEs have significant scientific, political, economic, and social impacts. A comprehensive framework is needed to guide and inspire an institution as a CoE and to help government and funding institutions shape and oversee CoEs.

**Strengths and limitations of this study:**

- To the best of our knowledge, this is the first scoping review to conceptualize centers of excellence based on global evidence.
- The study followed Arksey and O’Malley’s framework and methodological enhancement by Levac et al to and the PRISMA-ScR methodological frameworks.
- Five databases were systematically searched to identify scientific and gray literature
- The study was limited by language restrictions.

## Introduction

Globally, interest in excellence has grown exponentially, with public and private institutions shifting their attention from meeting targets to achieving excellence. The term center of excellence (CoE) has been understood as a team, shared facility, or entity that provides high standards of research, leadership, services or education, and brings innovative mechanisms to promote knowledge and scientific advancements (1-3). The concept has been around since copyrighted by Humana Health Plan (4); drawing the attention of scientists to enhance collaborations and cultivate access to resources essential for advanced research (5). It has been shaping institutions with ways to consolidate and build on their expertise and develop the capacity for scientific and financial gains (6). Organizations that work under the COE model have been maximizing the performance of their teams and nurturing productivity (7).

Nonetheless, there has been no consensus as to what CoE means and what operational modalities it should follow and ruled by (6,8). Although many centers are determined as CoE, there is no sufficient clarity as to what constitutes a COE, the resource requirements, and the processes and activities necessary to sustain the center. In the area of healthcare, CoE has been looked at as a specialized program with potential expertise and resources focusing on particular medical areas and providing exceptional outcomes (9, 10). Some experts in the field pinpoint particular fronts that distinguish CoE from traditional healthcare delivery models - organization design, servicescape design, personnel, medical care, marketing, and finance (9,11). Moreover, many research and higher education institutions (12-16) and healthcare institutions (17-20) across different countries have pursued a CoE status with claims that such institutions have bold distinctions in capacity, resources, and outcomes even among themselves.

Despite these assertions, there is ambiguity and inconsistency in the use of the term, the process of designations, processes of evaluation, and metrics. In this study, we conducted a global synthesis of the evidence to conceptualize CoEs.

## Methods

We followed a scoping review procedure to provide greater conceptual clarity and map CoE from heterogeneous sources. Scoping review is the most preferred approach to systematically identify and map key concepts, theories, and sources of emerging evidence and gaps in the research. We used Arksey and O’Malley’s framework for scoping reviews (21), which has been further enhanced by Levac *et al* (22), to analyze the evidence. We also used the PRISMA-ScR (Preferred Reporting Items for Systematic Reviews and Meta-Analyses extension for Scoping Reviews) (23) to guide the retrieval and inclusion of the evidence. The review was conducted following the stages described below:

### Stage 1: Identifying the research question

This stage involved in-depth discussion and consultation with the study team and study collaborators. The main research questions were the following:

> *How are CoEs defined?*
>
> *What are the building blocks of a CoE?*
>
> *What are the frameworks and functions of CoEs?*
>
> *What does it need to establish and sustain a CoE?*

### Stage 2: identifying relevant studies

#### Search strategy and sources

Relevant empirical and peer-reviewed literature were sourced from electronic databases and search engines including PubMed, Scopus, CINAHL, and Google Scholar up to 01 January 2021. Hand-searching of contents were conducted for key journals including the Journal of Excellence, International Journal of Excellence in Education, Journal on Centers for Teaching and Learning, International Journal of Business Excellence, Journal of Excellence in College Teaching, Equity and Excellence in Education, Excellence in Medical Education, Journal of Universal Excellence, Excellence International Journal of Education and Research, and Measuring Business Excellence.

To ensure that all relevant information were captured, we searched the Google engine for gray literature including conference proceedings, working papers, newsletters, business documents, presentations and reports, government documents, technical documents, white papers, policies, and bulletins. The references of included literature and documents were also hand-searched to get any additional evidence.

The databases were searched using both natural language and controlled vocabulary for “excellence” and the following terms within publication title and/or abstract: Centers of Excellence; Centers of Excellence for Education; Centers of Excellence for Research; Sustaining Centers of Excellence; Barriers to Achieving Excellence; Leadership for Excellence; Promoters of Excellence; Infrastructure for Excellence; and Collaborations for Excellence. The search term ‘‘Excellence’’ was further extended to include all relevant words through a search of definitions from dictionaries.

#### Eligibility

To be included in the review, sources of evidence needed to describe CoE as the main theme, which could be defining, theorizing, implementing, or evaluating a CoE. The search had no restrictions applied on the year of publication, publication status, or geographical location to ensure comprehensiveness of coverage, while non-English language literature were excluded. Duplicate citations were also excluded from the initial database.

### Stage 3: study selection

An extensive title screening, summary screening, and full-text screening strategy were developed to identify all relevant literature and documents. The title and abstract of all records retrieved were downloaded to and independently assessed for eligibility and screened by two authors after removal of duplicates. Disagreements between the reviewers were decided by consensus. Full-text copies of potentially relevant papers were retrieved for in-depth review.

### Stage 4: data charting

Data extraction used a standardized form developed by the investigator team. Information was extracted from each included document on 1) description of the document, including the primary author, publication year, and search engine; 2) Thematic area; 3) Essential foundations of the CoE, and 4) description of the major finding of the article. We checked if the CoEs had their status approved by an independent body or if they have a standard recognition to function as a CoE. We also checked if the CoEs have described the major barriers and opportunities in the establishment and sustaining of their CoE. Before extracting the actual data, the data extraction sheet was pilot-tested on 10 randomly selected studies, and the extraction sheet was refined as needed. Two authors (TM, AF) extracted the data.

### Stage 5: Collating, summarizing, and reporting the results

The results of the review were synthesized and narratively reported. The steps included thematic content analysis for qualitative information; numerical counts and tables for quantitative data; narrative summary and interpretation of all the results; and discussion of gaps in the literature.

Assessing the risk of bias across studies was not feasible due to expected high levels of variation in scope, setting, and time between studies. All given results were included to minimize the risk of selective reporting.

### Patient and Public Involvement

No patient involved.

## Results

### Study selection in the PRISMA review

Overall, 52,161 potential documents were included in the initial review from PubMed (n = 15,778), Scopus (n = 7,168), CINAHL (n = 1,150), Google Scholar (n = 10, 213), Google engine (n = 8, 142), and other sources (n = 23), of which 23,432 were retained after removing duplicates. Following an initial screening, 346 full-text articles were evaluated, of which 78 met the inclusion criteria and were included in the review (**Figure 1)**.

**Figure 1:**
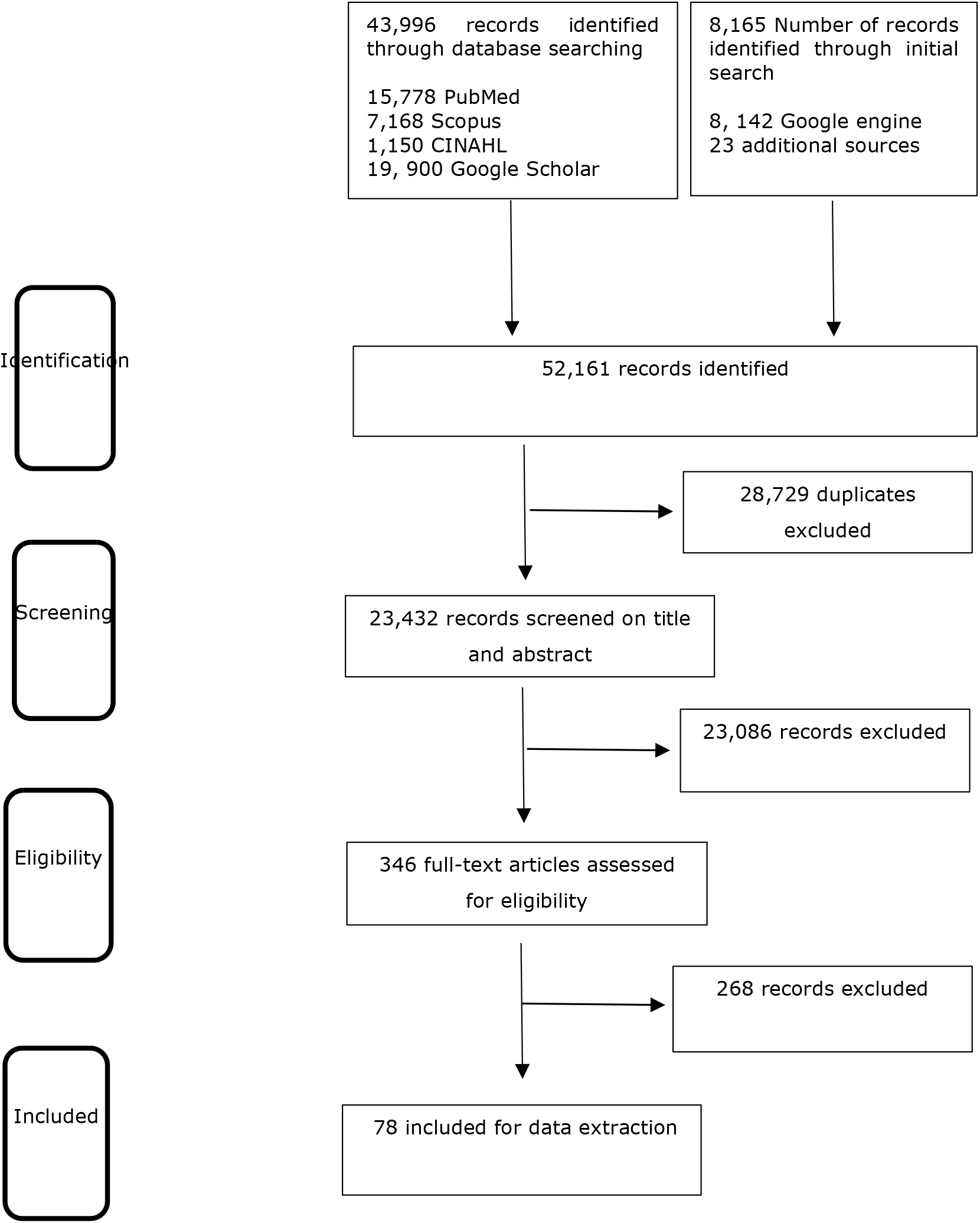
Flow diagram of the study

### Characteristics of the included studies

The 78 full-text articles included were affiliated with institutions globally from 33 countries, of which 35 were from the United States of America, 3 each from Nigeria, South Africa, Spain, and India, and 2 each from Ethiopia, Canada, Russia, Colombia, Sweden, Greece, and Peru. The rest 17 were from various countries.

The articles involved six thematic areas - health services, education, research, industry, information technology, and a general concept of CoE. **Table 1** summarizes the characteristics of included studies, including the name of the primary author and publication date, country of study origin, name of the designated CoE if any, thematic area of the literature, the essential foundation of the CoE, and a brief description of the key message in the literature.

**Table 1:** Characteristics of studies included in the scoping review

| 1<sup>st</sup> author, year | Country | CoE | Thematic area | Essential foundations of CoE | Description |
| --- | --- | --- | --- | --- | --- |
| Gao et al 2020 (24) | USA | US Food and Drug Administration (FDA) Oncology Center of Excellence | Health, Regulatory | Specialized expertise, Innovation, Collaboration | Targets to achieve patient-centered regulatory decision-making through innovation and collaboration |
| UTHealth 2020 (25) | USA | UTHealth COVID-19 Center of Excellence | Health care, Education, Research | Specialized expertise<br>Infrastructure<br>Advanced capacity | Established at the University of Texas Health Science Center at Houston (UTHealth) to lead the path forward in patient care and research |
| Bialous et al 2020 (26) | Czech Republic | Eastern Europe Nurses' Center of Excellence for Tobacco Control | Healthcare/ Public health, Education | Specialized expertise, Partnership<br>Educationally sound program | Address the epidemic of tobacco use and the resulting cancers in Central and Eastern Europe |
| Lipsitz et al 2020, (27) | Canada | Canadian Rapid Treatment Center of Excellence | Healthcare Research | Specialized expertise | It offers breakthrough rapid onset treatments for treatment-resistant conditions including depression and bipolar disorder |
| Komninaka et al 2020 (28) | Spain | Center of excellence on Rare Metabolic Disease- -Gaucher Disease | Healthcare Research | Specialized expertise | Treatment and follow-up of patients with Gaucher Disease |
| Araujo-Castro et al, 2020 (29) | Spain | Spanish Pituitary Tumor Center of Excellence | Healthcare Research | Specialized expertise (lead tea and supporting units) | It provides care delivery to patients with pituitary disorders |
| Balkhy et al 2020 (30) | USA | Robotic Heart Center of Excellence | Healthcare | Specialized expertise<br>Accreditation | Allina Health's United Hospital earned the accreditation and uses cutting-edge technology to provide the best and safest care. |
| Tince et al 2020 (31) | USA | Center of Excellence in Transgender Health | Healthcare | Organizational culture, Specialized expertise, Funding | Under the Gender Wellness Center, it fills gaps in rural transgender care |
| Sarkar et al, 2019 (32) | India | Center of Excellence in HIV care | Healthcare, Education, Research | Specialized expertise, High-impact research | Under the Calcutta School of Tropical Medicine in India, it provides care and conducts advanced research on HIV/HBV coinfection antiviral therapy. |
| Neal et al 2018 (33) | Rwanda | Butaro Cancer Center of Excellence | Healthcare | Specialized expertise, Facility | It offers a spectrum of diagnostic oncology and treatment services. |
| Choque-Velasquez et al 2017 (34) | Peru | Neurosurgical Center of Excellence | Healthcare | Specialized expertise, Facility | Under the EsSalud hospital of Trujillo, it works to improve the treatment of neurosurgical diseases in the local region |
| Elrod et al, 2017 (35) | USA | Concept on CoE in healthcare institutions | Healthcare | Specialized expertise, Resources, Understanding delivery model | The authors defined COE in healthcare and described what they are and how to assemble them. |
| Santos-Moreno et al, 2017 (36) | Colombia | centers of Excellence for Rheumatoid Arthritis | Healthcare | Specialized expertise, Standards | The authors proposed a systematic and progressive methodology that will help all the institutions to develop successful models for CoE in rheumatoid Arthritis. |
| Chick et al, 2017, USA (37) | USA | Hereditary Hemorrhagic Telangiectasia Centers of Excellence | Healthcare | Specialized expertise | It proactively provides care for patients with Hereditary Hemorrhagic Telangiectasia. |
| García-Perdomo, 2017 (38) | Colombia |  |  | A multidisciplinary board and expertise, Special care units, Accreditation, Electronic Medical Record System, Leadership | The author underlined a COE is not any healthcare facility that receives and attend people with a determined condition, but one that needs to fulfill certain requirements. |
| Nwafor, 2017 (39) | Nigeria | National Cardiothoracic Center of Excellence | Healthcare Research | Collaboration and partnership | Under the University of Nigeria Teaching Hospital, the COE provides cardiothoracic services |
| Casanueva, et al, 2017 (40) | Spain | Pituitary Tumor Centers of Excellence | Healthcare | Specialized expertise, Leadership, Organizational structure | The authors generated criteria for developing Pituitary Tumors COE to guide improvements of care delivery to patients with pituitary disorders |
| Cohen, 2017 (41) | USA | Concept on CoE | Healthcare Education, Research/scholarship | Healthcare expertise, Innovation Entrepreneurship | The authors described the process that the University of Michigan created as the first pathway of excellence for medical students to focus their passions and interests in medical innovation and entrepreneurship. |
| Doumouras et al, 2017 (42) | Canada | Regionalized Center of Excellence Bariatric Care System | Healthcare | Healthcare expertise, Accreditation | The COE provides obesity treatment, assesses and treats patients by following evidenced-based strategies, standards, and protocols recommended by the Ontario Bariatric Network (OBN) and approved by the Ministry of Health and Long-Term Care. |
| Scheuermann, et al, 2017 (43) | USA | Centers for Quantitative Imaging Excellence | Regulatory | Accreditation | The COE prequalifies imaging facilities at all of the National Cancer Institute-designated comprehensive and clinical cancer centers for oncology trials using advanced imaging techniques, including PET. |
| Miranda et al, 2016 (44) | Peru | CRONICAS Centre of Excellence in Chronic Diseases | Healthcare Research | Collaboration and partnership | Universidad Peruana Cayetano Heredia, the COE fosters sustainable partnerships for international collaborative research and improves population's health through high-quality research |
| Kouchoukos, 2016 USA (45) | USA | The concept on Cardiothoracic Surgical Center of Excellence | Healthcare | Accreditation Standards | The author describes the term COE is used without official designation, specific or measurable criteria, and external review. Providers of cardiothoracic surgical care should refrain from referring to their organizations as COEs until specific criteria are established by the appropriate specialty societies. |
| Melissas et al 2017 (46) | Greece | IFSO-European Chapter Center of Excellence | Healthcare | Accreditation Training/ Education | The COE is evolved from the rapid growth of the field of obesity, started by a group of surgeons and grown to become a world organization. It provides accreditation of individual surgeons and bariatric centers around the globe. |
| Singh et al, 2015 (47) | India | AIIMS Neurosciences Center of Excellence | Healthcare Research Education | Expertise, Collaboration and partnership, Infrastructure | Under the All India Institute of Medical Sciences (AIIMS) the COE works to the advancement and promotion of neurosciences in India |
| Rábago, 2016 (48) | USA | Extremity Trauma and Amputation Center of Excellence | Healthcare Research | Specialized expertise, Innovation, Leadership, Collaboration and partnership | Under the Department of Defense and Department of Veterans Affairs, the COE is mandated to optimize the quality of life of service members and veterans who sustain extremity trauma or amputation, and research novel rehabilitation interventions, advanced prosthetic and orthotic technologies, epidemiology and surveillance, and medical and surgical innovations. |
| Rani, 2016 (49) | India | Retinopathy of Prematurity (ROP) Tertiary Centers of Excellence | Healthcare Education/ training | Specialized expertise, Funding, Infrastructure Leadership | The COE provides integrated capacity building, education, training and awareness-creation program to enhance ROP screening. |
| Mettrick et al, 2015 (50) | USA | Concept on COE development, with emphasis on COE for child serving agencies | Cross-cutting | Infrastructure, Partnership, Policy and financing support, Subject matter expertise | The authors proposed a roadmap for state and local child-serving agencies in developing COEs; and defined COE from different perspectives |
| Harefuah 2015 (51) | Israel | Meir Medical Center of excellence in care quality, service and research | Healthcare Research | Infrastructure, Long-term strategy, Leadership, Scientific community | The Center achieved COE through initiatives including research grants, scientific meetings, development of infrastructure, long term strategy for academic research. |
| Mehrotra et al, 2015 (52) | USA | Concept on COE – how to ensure excellence in centers of excellence programs. | Healthcare | Structural and process of care, Accreditation/ formal designation | The authors underlined, the outcomes at COE must be clearly superior to ordinary institutions to outweigh these potential harms, and that moving forward, there is a need to change how hospitals are designated and most importantly provide evidence that the COEs are truly excellent. |
| Katz et al, 2015 (53) | South Africa | Centre of Excellence in HIV care | Healthcare | Quality of care - social support, adequate preparation for transfer, and improving the quality of care | A community-based clinics from Sinikithemba was considered a Centre of Excellence in HIV care, at McCord Hospital, with PEPFAR-associated transitions. |
| Itri et al, 2014 (54) | USA | Concept on COE – How the Six Sigma used to improve Outpatient center of excellence in CT | Healthcare | Conformance to requirements. Improve patient experience, Improve staff satisfaction | The authors described how to Create an outpatient COE in CT to improve patient experience and increase CT capacity while reducing waste and improving staff satisfaction. |
| Reichert et al, 2014 (55) | USA | Concept on COE - 5 key pillars of an analytics COE, | Healthcare | Governance, Organizational, Structure, People, Process, Technology, Institutional culture | The authors identified 5 key pillars of a COE required to manage populations and transform organizations into the next era of health care |
| Zar et al, 2014 (56) | South Africa | Concept on COE - Partnering with COE in high- and low–middle-income countries | Healthcare | Partnership | Partnerships need to be undertaken in a mutually beneficial, collaborative, and respectful way; LMICs need to identify their needs for expertise and professionals in high-income countries need to undertake such collaborations with the goals of developing and building local capacity in low- and middle-income settings and of transferring technology and skills |
| Ferguson et al, 2013 (57) | USA | Establishing a Center of Excellence | Healthcare | Collaboration Leadership | At The Miriam Hospital, The Total Joint Center has been very successful with the cooperative relationship between various stakeholders. |
| Mehrotra et al, 2013 (58) | USA | Center of Excellence Program for Spine Surgery | Healthcare | Empirical evaluation | Assessing whether hospitals designated as spine surgery COE by a group of over 25 health plans provided higher quality care, the authors reported spine surgery CoEs had similar complication and readmission rates compared to other ordinary hospitals. |
| NIH, 2009 (59) | USA | NIH centers of excellence | Healthcare Education Research | Infrastructure, Collaboration, Training | The COEs' programs help establish critical research infrastructure; foster collaboration; train researchers, physician-scientists, and other professional staff; and provide shared resources, often through core facilities. |
| Sharkey et al, 2009 (60) | USA | Concept on COE - Challenges in sustaining excellence | Healthcare | Infrastructure | Based on their experience at Saint Joseph's Hospital of Atlanta, the authors underlined a culture of excellence is not synonymous with a culture of perfection as perfection is not attainable but excellence is when infrastructure is developed that can rapidly and effectively adapt to change. |
| Cullen, 2005 (61) | USA | Concept on COE - Excellence in Evidence-Based Practice | Healthcare | Capacity, Culture, Vision of the organization | Creating excellence begins by building the capacity, culture, and vision for the organization and the unit. The organization builds a capacity for evidence-based practice when developing the infrastructure |
| Keto et al, 2007 (62) | USA | Centers for Medicare and Medicaid Services | Healthcare | Specialized expertise, Infrastructure and equipment, Leadership, | The Centers for Medicare and Medicaid Services requirements for Centers of Excellence designation resulted in a significant increase in the Medicare caseload. |
| Anderson et al, 2002 (63) | USA | National Centers of Excellence in Women's Health | Healthcare | Quality of care, Patient satisfaction | There was improved quality of care in the Department of Health and Human Service (DHHS)-designated CoEs compared with national and local community benchmark samples. |
| Pennsylvania DHS (64) | USA | COE on addiction | Healthcare | Regulatory, Funding, | Pennsylvania Department of Human Services (DHS) funds providers who are interested in becoming COE – that has health homes, coordinate care for people with Medicaid. |
| Dieye et al, 2016 (65) | Nigeria | West Africa International Centers of Excellence for Malaria Research | Research | Collaboration | The COE is a Multidisciplinary Research for Malaria Control and Prevention in West Africa with 4 research focus: Malaria Epidemiology, Immuno-Genetics, Malaria Vector Ecology, and Supplement Projects. |
| Nwaka et al, 2012 (66) | Ethiopia | pan-African Centres of excellence in health innovation | Research | Innovation, Collaboration and networking, Specialized expertise. | African Network for Drugs and Diagnostics Innovation (ANDI) been selected and recognized 38 pan-African CoEs in health innovation through competitive criteria. |
| Fekadu et al, 2014 (67) | Ethiopia | Center for Innovative Drug Development and Therapeutic Trials | Research, Education | Funding, Partnership, Infrastructure, Innovation | Under the Addis Ababa University, an Africa center of excellence is established to build regional capacity for translational research and clinical trials. |
| Mccarthy 2017 (68) | France | ESRF: Excellence in Service to Users | Research | Partnership, Innovation | The ESRF is a COE for fundamental and innovation-driven research in condensed and living matter science. |
| Heining et al, 2016 (69) | German | Oncological centers of excellence | Healthcare Research | Infrastructure, Specialized expertise, Funding | German Cancer Aid launched an oncological COE program and provides funding for 13 CoEs that have to fulfill defined criteria and undergo periodic reevaluation and certification by international experts. |
| Bloch et al, 2016 (70) | Denmark | Grant size for Centres of Excellence | Research | Funding size, Funding period | Performance for these CoEs funded by the Danish National Research Foundation has been very high, with performance increasing in grant size and over time. |
| Borlaug, 2016 (71) | Norway | Centres of excellence in Norway and Sweden | Research | Funding | in a country with a highly competitive funding system, funding agencies emphasize organizational impact to overcome the the problem of moral hazard. |
| Pislyakov et al, 2014 (72) | Russia | Russian centers of excellence | Research | Collaboration, High-impact research | More than 90% of Russian highly cited papers from CEOs involve international collaboration, and Russian institutions often do not play a dominant role. |
| Househ et al, (73) | Saudi Arabia | Electronic Health Center of Research Excellence (E_CORE) | Research Electronic health | Innovation, Quality of care, Cost-effectiveness | ECORE is established to become a world leader in e-health research and training through innovation and research excellence, improving healthcare quality, and reducing healthcare costs. |
| Lin et al, 2009 (74) | Taiwan | National Center of Excellence for Clinical investigators and research | Research | High-impact research, Leadership in clinical research, Networking | At the National Taiwan University Hospital, the COE, focuses on developing new treatments for endemic diseases in Taiwan through clinical trials focusing on targeted therapy. |
| Rocco et al, 2015 (75) | Italy | Center of Excellence to Build Nursing Scholarship | Education Healthcare | Funding, Collaboration | The concept of COE is a potential instrument to strengthen nursing scholarship in Italy |
| Fetrow et al 2020 (76) | USA | National Center of Excellence in Dairy Production Medicine Education for Veterinarians | Education | Innovation | The authors described an effort to create a dairy production medicine curriculum funded by a United States Department of Agriculture Higher Education Challenge Grant |
| Harris et al 2020 (77) | USA | Center of Excellence in Health Equity, Training and Research | Education, Research | Innovation cutting-edge ideas, Partnership | At Baylor College of Medicine, the COE organizes Health Equity Summer Research Summit to catalyze the fertilization and exchange of cutting-edge ideas in the area of disparity research in medicine. |
| World Bank 2021 (78) | USA | Eastern and Southern Africa Higher Education Centers of Excellence | Education and research in multiple priority areas | Collaboration, Leadership, Strategy | Supported by the World Bank, the project strengthen selected Eastern and Southern African higher education institutions to deliver quality postgraduate education and build collaborative research capacity in the regional priority areas. |
| Nwafor et al 2017 (79) | Nigeria | National Cardiothoracic Center of Excellence | Education Healthcare | Collaboration | University of Nigeria Teaching Hospital, the COE, through the aid of visiting Cardiac Missions, has managed a significant number of patients. |
| Kumar et al 2017 (80) | USA | MD2K: Center of Excellence for Mobile Sensor Data-to-Knowledge | Health, Research, Education | Innovation, Funding | The COE was established with funding from the NIH under the Big Data-to-Knowledge program to develop innovative tools to make it easier to gather, analyze, interpret, and capitalize on high-frequency data from mobile sensors. |
| Giacomini et al 2019 (81) | USA | University of California, San Francisco-Stanford Center of Excellence in Regulatory Science and Innovation | Regulatory science, Research, Education | Innovation Partnership, Scientific exchange | The COE promotes regulatory science—including innovative research, education, and scientific exchange—in partnership with foundations and commercial entities interested in the development of FDA-approved medical products. |
| Terama et al, 2016 (82) | UK | Concept on COE - Interrogating Research Impact in the Research Excellence Framework | Research | High-impact research | The authors found a positive correlation between impact score and overall research quality score, suggesting that the impact is not being achieved at the expense of research excellence. |
| Cofrancesco, 2017 (83) | USA | The Institute for Excellence in Education (IEE) | Education | Innovation, Funding, Partnership | Key to the IEE's success in maintaining and building programs has been ongoing needs assessment of faculty and learners and a strong partnership with fund-raising staff. |
| Voznesenskiy et al, 2016 (84) | Russia | ITMO Photonics: center of excellence | Research, Education | Infrastructure, Specialized expertise, Strong financial base | At ITMO University, the ITMO Photonics COE disseminates breakthrough scientific results in photonics into an undergraduate and graduate educational program, concentrating organizational, scientific, educational, financial, laboratory and human resources. |
| Bienenstock et al, 2014 (85) | Sweden | Combining excellence in education, research and impact | Education, Research | High-quality teaching, Interdisciplinary skills | The authors compared Swedish universities with Berkeley and Stanford for key determinants of universities' excellence, relevance and global competitiveness, and found significantly lower quality and focus of teaching and education with Swedish universities. |
| Rugen et al, 2014 (86) | USA | Centers of Excellence in Primary Care Education | Education | Innovation, Partnership, Collaboration, Quality improvement | The Department of Veterans Affairs has funded five CoEs in Primary Care Education to help develop and test innovative structural and curricular models. |
| Bittner et al, 2013 (87) | USA | Concept on CoE – CoE as a designation | Education | High-quality education | At Regis College, a COE designation in student learning and professional development enable nursing students to excel academically, professionally, and personally. |
| Balaji, 2017 (88) | Malaysia | Marine engineering center of excellence | Education | Infrastructure | For building an engineering center for training, mock-up facility, simulator, full-scale engine room, scaled-down version and a combination arrangement with live and dummy equipment were options at different physical levels. |
| Sobhieh, 2016 (89) | Iran | COE at power plant projects | Industry | Skilled expertise | Establishing CoE can be effective in achieving human capital in Iran. |
| Geiger, 2006 (90) | USA | Concept on CoE – Establishing COE | Industry | Collaboration, Best practice | CoE is a team of people that is established to promote collaboration and the application of best practices and has three characteristics: authority, role, and organization placement and staffing |
| Nqampoyi et al 2016 (91) | South Africa | Concept on CoE - Business Process Management Centres of Excellence | Industry | Standard, Service quality and scope | The services and industry standards have the largest impact on the effectiveness of the CoE. |
| Neely et al, 2017 (92) | Sierra | IT application center of excellence | Information Technology | Specialized expertise, Collaboration | Collaboration with strong companies ensures a successful transition of the COE's mission-oriented applications into the exascale era. |
| Wang, 2017 (93) | China | CAS Center for Excellence in Quantum Information and Quantum Physics | Information Technology | Innovation | Under the China Academy of Sciences (CAS), the COE overcome the bottleneck of classical IT in ensuring information security, enhancing computing capacity, and improving measurement precision. |
| Craig et al, 2009 (94) | USA | Concept on CoE - Criteria and evaluation method for COE | Information Technology | Internal business process, Customer focus Leadership, Innovation and learning, Finance | Understanding the gaps in how CoEs are designated, accredited, or certified and the means of auditing, assessing, or appraising them, the authors proposed criteria and standards to certify an organization as a CoE are presented in the Carnegie Mellon Software Engineering Institute. |
| Grant et al, 2006 (95) | USA | Concept on CoE – Measuring excellence | Cross-cutting | Leadership, Best performance, Customer satisfaction | Excellence is more than doing better than average and is all about outperforming by a substantial margin |
| Patel et al, 2010 (96) | USA | Concept on COE - Steps to establishing A center of excellence | Cross-cutting | Organizational structure, Guiding principles, Scope | A charter for the CoE must be defined, including its vision statement, guiding principles, and scope with three types - knowledge-sharing, strategy/guidance, and implementation/support. |
| Marciniak, 2012 (97) | Hungary | Concept on COE - COE as a next step for shared service center | Cross-cutting | Leadership, Organizational efficiency | There are more success stories about the potential of COE in improving organizational efficiency. A long-term, comprehensive cost-conscious business plan transforms a profit center from a cost center. |
| Hellström, 2013 (98) | Sweden | Concept on COE - COE as a Tool for capacity building | Cross-cutting | Innovation, High impact research, Specialized expertise | The author categorizes CoE schemes into basic and strategic research; innovation and advanced technological development; and social and economic development. CoEs are considered organizational environments that strive for and succeed in developing high standards of conduct in a field of research, innovation, or learning. |
| Brusoni et al, 2014 (99) | Belgium | Concept on COE – the concept of COE in higher education | Education | Accreditation or standard, Quality improvement, | The approach to excellence is progressive and not all agencies recognize excellence. Specification of excellence provides a framework for a focus for enhancement. |
| Heyes, 2012 (100) | USA | Nuclear Security Centers of Excellence | Nuclear security | Technical and scientific expertise, Global partnership, Coordination and collaboration | Based on an assessment of the currently active COEs in the area, and those planned for implementation, the author categorized the COEs into five groups. |
| Antonowicz et al, 2017 (101) | Poland | Concept on COE - The roads of 'excellence' in Central and Eastern Europe | Education | Quality assurance, political drivers, Economic mechanisms, Institutionalization | The academic community understands and shares enthusiasm towards excellence, but it also shows some resistance towards making it a top political priority, which legitimizes rapid and deep structural changes in higher education. |

### Definition of Center of Excellence

CoE is a team of specialized expertise (25-37, 40-42, 47, 49, 62, 66, 69, 84, 85, 89, 92, 98, 100) or organizational environment (25, 31, 35, 38, 40, 41, 50, 51, 55, 61, 74, 94, 96, 97, 101) that is established to provide outstanding healthcare (24-42, 44-49, 51-64, 69, 73, 75, 79-80), research (25, 27-29, 32, 39, 41, 44, 47, 51, 59, 65-74, 77, 78, 80-83), education and training (25, 26, 32, 41, 47, 49, 59, 67, 75, 77-81, 83, 84-88), regulatory (24, 43, 81), policy (50, 78, 95-98), information technology (92-94) or industrial (89-91) services and support in high levels of efficient and effective performance. CoEs are geographically concentrated and focused on high potential in science and industry (98), as a world leader or a catalyst between neighboring countries (46, 73, 78), anywhere from the local R&D group up to a network of co-operative partners. They are characterized by the scope of their operations, mandates, funding, executive sponsorship, commitment, responsibilities, and powers (99). Historically, CoE is leveraged by IT leaders seeking to facilitate the creation of hubs for knowledge sharing and building and enhancing capabilities. The idea had evolved and been used in favor of different targets in the last decade (97). Its driver, “Excellence”, was not considered an act that has to do with the end product, but a habit to do with the process.

### Functions of Centers of excellence

When CoEs are established, it is most often because of a need for implementation infrastructure, existence of specific relationships, and availability of funding (50). Its establishment could favor critical research infrastructure, foster collaboration, train experts, and share core facilities (59). Regardless of strategic orientation, all CoEs have in common the notion of excellence and the particular requirements that come with that label, such as high research quality and productivity, resource attraction and concentration, international visibility and attractiveness, and organizational robustness (98).

Within the vertically different higher education, excellence is being equated to ‘being better’ which could mean, excellence in research, top-quality professors, favorable working conditions, job security and good salary and benefits, adequate facilities, adequate funding, academic freedom, an atmosphere of intellectual excitement, and faculty self-governance (99). Higher education institutions develop such CoEs to address the ongoing critical need for more intensive and specialized training on a specific condition (76, 77, 79).

In the healthcare sector, CoEs can provide breakthrough treatments on specific disease conditions (31, 27-30, 32-34). In line with the current COVID-19 pandemic, some hospitals and universities have established COVID-19-specific CoES [25, 102].

CoE could be established to provide specific regulatory decision-making, including to advance the development and regulation of products [26, 81] and to fill-in knowledge gaps [103]. The world Bank has an Eastern and Southern Africa Higher Education Centers of Excellence project that strengthen selected Eastern and Southern African higher education institutions to deliver quality postgraduate education and build collaborative research capacity in the regional priority areas (78).

### Essential foundations of Center of Excellence

There were 12 essential foundations of CoE according to the review of the 78 literature (**Figure 2**).

**Figure 2:**
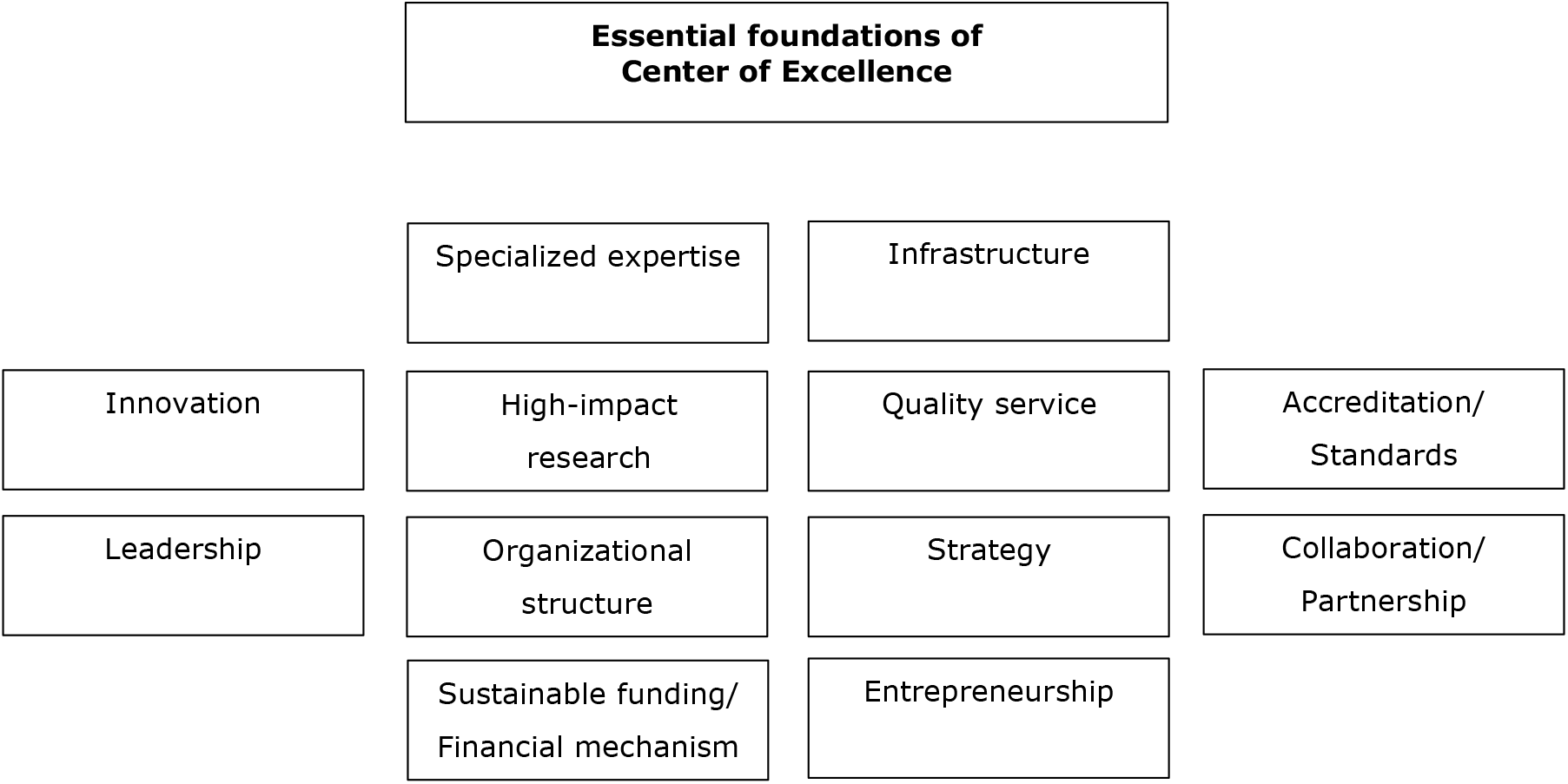
Essential foundations of CoE in literature

### Frameworks and models of Center of Excellence

Fourteen of the 78 included articles have used or developed a specific framework or model to establish and shape their institutions as a CoE or to distinguish CoE from the traditional institutional or service delivery model. **Table 2** summarizes the frameworks employed.

**Table 2:** Frameworks and models of Centers of Excellence

| Framework or model | Literature that cited the framework/model |
| --- | --- |
| Willis-Knighton's Health System CoE delivery model | Elrod et al, 2017 (35) |
| Lean Six Sigma | Itri et al, 2014 (54); Ferguson et al, 2013 (57) |
| REAL-PANLAR Group quality standard requirements for CoE model | Santos-Moreno et al, 2017 (36) |
| Curriculum development framework | Cofrancesco, 2017 (83) |
| Four-domain curricular model | Rugen et al, 2014 (86) |
| Research Excellence Framework | Terama et al, 2016 (82) |
| IFSO requirement for CoE program participation | Melissas et al, 2016, Greek (46) |
| Three-impact dimensions | Borlaug, 2016 (71) |
| Key steps to establishing a CoE | Patel et al, 2010 (96) |
| Generalized Criteria and Evaluation Method for CoE | Craig et al, 2009 (94) |
| Excellence in higher education evaluation model | Brusoni et al, 2014 (99) |
| E-CoRE Conceptual framework | Househ et al, (73) |
| Critical capacities of a CoE | Mettrick et al, 2015 (50) |
| Activity-based typing of CoEs | Heyes, 2012, (100) |

Elroad et al (35) looked at CoE under the umbrella of the Willis-Knighton Health System and identified six particular fronts - organization design, servicescape design, personnel, medical care, marketing, and finance - that distinguish CoE from traditional healthcare delivery models. Addressing each of these fronts yield an exceptionally high level of care largely exceeding that delivered in traditional settings. Elroad et al considered CoE in healthcare as a program that is assembled to supply an exceptionally high concentration of expertise and related resources centered on a particular area of medicine, delivering associated care in a comprehensive, interdisciplinary fashion to afford the best patient outcomes possible.

Itri et al (54) and Ferguson et al (57) used the Lean Six Sigma framework with its five distinct phases - define, measure, analyze, improve, and control - to establish CoE in healthcare settings. The authors proved that the framework is effective in establishing and organizing CoEs.

Santos-Moreno et al (36) employed the REAL-PANLAR Group model as a requirement for implementation and accreditation of COEs in rheumatoid arthritis care. The framework defined three types of CoEs based on structure, process, and outcome indicators - standard, optimal, and model. To maintain the comprehensive nature of care, all criteria must be met with a minimum score of 80% for each of the criteria. The approved standards for the “standard” CoE should have a basic structure and process indicators, the “optimal” level should accomplish more structure and process indicators, and the “model” level should fulfill outcome and patient experience indicators. In addition and regardless of institutional category, all CoEs should meet the same standards, and according to the philosophy of the accreditation system, the concepts of continuous improvement and patient-centered management must be included.

Cofrancesco et al (83) used the six-step curriculum development framework to inspire and support the development and outcomes of excellence in education programs. The steps included problem identification and general needs assessment; targeted needs assessment; setting goals and objectives; educational strategies; program implementation; and evaluation and feedback. The authors developed and implemented the Institute for Excellence in education as a structure that works to promote, improve, and innovate medical and biomedical education and scholarship.

Rugen et al (86) developed a curricular model with four domains - shared decision making, sustained relationships, inter-professional collaboration, and performance improvement – aimed at achieving CoE in primary care education. The model transformed health care training from a profession-specific primary care delivery approach to an inter-professional, team-based, patient-centered care delivery paradigm.

Terama et al (82) applied Research Excellence Framework to assess the quality and reach of research in UK universities and allocate funding accordingly. The framework assesses the quality of research in higher education institutions in terms of three elements - originality, significance, and rigor of research outputs; reach and significance of impact; and vitality and sustainability of research environment. The framework was instrumental in establishing education excellence, while it was unlikely to fully reflect all of the impacts of university research.

Melissas et al (46) applied the IFSO (Surgery of Obesity and Metabolic Disorders) requirement for CoE program participation. The IFSO provides designation as CoE in bariatric and metabolic surgery for institutions fulfilling ten requirements focusing on experts’ capacity, infrastructure, a facility set up, and appropriate technologies. The authors improved healthcare services following participation in the IFSO.

Borlauge (71) developed a conceptual framework involving three impact dimensions - organizational, social, and international – to examine how CoE schemes have been adapted to two distinct national public research systems in Norway and Sweden. With this framework, the author identified that funding agencies emphasized organizational impact in a country with a highly competitive funding system.

Patel et al (96) proposed a seven key-steps framework to consider in creating a CoE. The steps included defining the CoE and setting vision statement and guiding principles, determining the overall scope for the COE which has three types including knowledge-sharing focus, strategy/guidance focus, and strategy/guidance focus with implementation/support focus.

Craig et al (94) proposed a framework that characterized the successes of a CoE through five dimensions - internal business process, customer focus, leadership, innovation and learning, and finance. The framework integrated elements of the balanced scorecard and the Baldrige Criteria for Performance Excellence to form a background for evaluating organizations as CoE.

Brusoni et al (99) framed seven evaluation extents to define excellence in higher education – leadership, purposes and plans, beneficiaries and constituencies, programs and services, faculty/staff and workplace, assessment and information use, and outcomes and achievements. Excellence in Higher Education provides a structured guide for reviewing each of these areas as they operate within a particular institution, department, or program.

Househ et al (73) were guided by a conceptual framework that is comprised of the production, dissemination, and use of knowledge in their establishment of an Electronic Health Center of Research Excellence (E-CoRE). In the framework, knowledge was produced as a result of research efforts, innovation and collaboration, and such generated information is used to formulate decisions.

Mettrick et al (50) indicated that every COE’s scope of work should have four critical capacities - implementation support, continuous quality improvement for identified interventions, workforce development, and technical assistance. As the initial scope of work is refined, these criteria should be used to begin to identify possible organizational candidates to house the CoE.

Heyes (105), as part of his assessment of the nuclear security CoEs, grouped CoEs into five types based on their core activities. Group A Centers where the core activities are essentially technical and scientific with a focus on providing training on the use, calibration, and maintenance of equipment. Group B Centers where the core activities are essentially educational, offering the course(s) which, although they may have technical content, are designed to provide a broad perspective of nuclear security and an awareness of relevant issues. Group C Centers where the core activities encompass a wider range of topics than just nuclear security, or even wider than nuclear security, safety, and safeguards. Group D Centers where the core activity is focused on nuclear research and development or which are characterized by strong commercially driven objectives. Group E Centers where the core activities are focused on raising awareness of nuclear security issues within the nuclear industry and beyond.

## Discussion

Our analysis of the scientific and gray literature documented success stories of using the brand “CoE” within the healthcare, research, education, information technology, and industry; signifying the huge potential of functioning as a CoE. There were some key milestones commonly applicable to many CoEs, despite the inconsistent use and self-designation of the brand “CoE” without approval by an independent, external process of evaluation, and with high ambiguity between “CoE” and the ordinary “Institution” or “Center”. In the developing world, the concept of CoE is getting significant attention, particularly in research and higher education sectors where senior professors are concentrated, as our analysis pinpoints. Our review led us to define CoE as a team of specialized expertise or organizational environment that is established to provide outstanding healthcare, research, education and training, regulatory, information technology or industrial services, and support in high levels of efficient and effective performance.

Our analysis showed that a comprehensive framework is needed to guide and inspire an institution as a CoE. There were some existing frameworks that the CoEs used as a benchmark, to shape the establishment and implementation of their CoEs; however, there were some important limitations with the frameworks:

1. The frameworks were limited to demarcating the unique features of a CoE as compared to an ordinary institution.
2. The frameworks were limited to specific facilities or services without analysis at a larger scale.
3. The frameworks lacked parameters and levels of indicators to use to follow-up or improve the performance of the CoEs, unlike many accreditation requirements for different research and education activities.
4. The frameworks were not sufficient to enhance CoEs to acquire excellence in both research and education when these two activities are combined in a CoE, though there were many CoEs tagged as CoE for Research and Education. However, as one compliments the other it could be possible to achieve excellence in both if one exploits all the available opportunities in both areas.
5. Giving particular emphasis to CoEs in developing countries, the frameworks did not recognize the issue of resource potential and stewardships in such settings.

Nevertheless, the scoping review indicates that highly specialized expertise and infrastructure are the basic determinants of a CoE, with the number and multidisciplinary nature of the experts influencing the way the CoE functions and competes with the rapidly changing knowledge and technology landscape.

The CoE needs to ensure that it has the right team in place to achieve its vision and mission. The appointment of each of the team members needs to be merit-based, taking into account each member’s education, training, experience, and the broader vision of long-term requirements for the CoE. The CoE needs to maintain high efficiency and effectiveness distinctive from ordinary healthcare, research or education programs.

The CoE needs to have the quality infrastructure and technology, facilities, resources, and related services to conduct or facilitate research, and with capacity for specialized expertise from different contexts and countries to visit, practice, and collaborate. Structurally, the CoE’s could be a stand-alone promise or a confirmed part of a legal institution as a separate division in an academic or research institution, or a hub location for multi-country functions. The CoE needs to promote a global view aimed at enhancing healthy competitions on a global platform and promoting the development of globalized knowledge. It should benchmark its performance against equivalent CoEs in other countries and give sufficient attention to collaboration and joint works. Its researches should have a high-impact when compared to ordinary research institutions and they should have regional and international visibility attracting partnership, collaboration, and knowledge-transfer opportunities. It needs to develop its strategic direction with a broader vision, mission, goals, and objectives.

Leadership is one of the critical structures of a CoE in this regard. The center leadership should have the maximum commitment to achieving excellence, with the potential to influence the overall functions and long-term visions of the CoE. The leadership should explore sustainable financing mechanisms which could be sourced from government institutions, funding agencies, public-private partnership, or generation of revenues through consultancy, training or research services. The leadership should also make sure that its staff is highly motivated and ambitious.

## Conclusion

CoEs have significant scientific, political, economic, and social impacts. However, there are inconsistent use and self-designation of the brand without approval by an independent, external process of evaluation, and with high ambiguity between “CoEs” and the ordinary “Institutions” or “Centers”. A comprehensive framework is needed to guide and inspire an institution as a CoE and to help government and funding institutions shape and oversee CoEs.

## Data Availability

Not applicable.

## Declarations

### Authors’ contributions

Conceptualization, methodology, and first draft: TM, AF. Revised the draft: YW, CO, AH, AB, MG, GM, GY, AC, EM. All authors have read, reviewed, and approved the final manuscript for publication.

## Acknowledgments

The World Bank financially supports the Center for Innovative Drug Development and Therapeutic Trials for Africa (CDT-Africa).

## Availability of data and materials

Not applicable.

## Ethics approval and consent to participate

Not applicable.

## Consent for publication

Not applicable.

## References

1. Fekadu A, Oppenheim C, Manyazewal T, et al. Understanding the key processes of excellence as a prerequisite to establishing academic centres of excellence in Africa. BMC Med Educ. 2021;21(1):36.

2. Elrod JK, Fortenberry JL Jr. Centers of excellence in healthcare institutions: what they are and how to assemble them. BMC Health Serv Res. 2017;17(Suppl 1):425.

3. Nakov R, Sarafov S, Gospodinova M, et al. Transthyretin amyloidosis: Testing strategies and model for center of excellence support. Clin Chim Acta. 2020;509:228–234.

4. Itri JN, Bakow E, Woods J. Creating an outpatient center of excellence in CT. J Am Coll Radiol. 2014;11(12 Pt A):1137–1143.

5. Reichert J, Furlong G. Five key pillars of an analytics center of excellence, which are required to manage populations and transform organizations into the next era of health care. Nurs Adm Q. 2014;38(2):159–165.

6. Blokdyk G. Center of Excellence a Complete Guide, 2019 Ed. Australia; Emereo Pty Limited, 2019.

7. Santos-Moreno P, Caballero-Uribe CV, Massardo ML, et al. Systematic and progressive implementation of the centers of excellence for rheumatoid arthritis: a methodological proposal. Clin Rheumatol. 2017;36(12):2855–2858.

8. Walshe R, Herrmann-Frank A, Diehl V. Center of Excellence, Cancer Center, Kompetenznetz: Terminologischer Wirrwarr? [Center of excellence, cancer center, competence network: confusion of terminology?]. Dtsch Med Wochenschr. 2002;127(17):913–914.

9. FDA Compounding Quality Center of Excellence. J Nucl Med. 2020;61(3):21N–22N.

10. Drake J, Schreiber KC, Lopez R, et al. Establishing a center of excellence for hereditary diffuse gastric cancer syndrome. J Surg Oncol. 2019;119(6):673–674.

11. El-Eshmawi A, Castillo JG, Tang GHL, Adams DH. Developing a mitral valve center of excellence. Curr Opin Cardiol. 2018;33(2):155–161.

12. Nwafor IA, Chinawa JM, Adiele DK, et al. Management of complex CHD at the National Cardiothoracic Center of Excellence, University of Nigeria Teaching Hospital, Enugu: the role of foreign cardiac missions in 3.5 years. Cardiol Young. 2017;27(6):1174–1179.

13. De Angelis F, Abdelgawad M, Rizzello M, Mattia C, Silecchia G. Perioperative hemorrhagic complications after laparoscopic sleeve gastrectomy: four-year experience of a bariatric center of excellence. Surg Endosc. 2017;31(9):3547–3551.

14. Sejdić E, Rothfuss MA, Stachel JR, et al. Innovation and translation efforts in wireless medical connectivity, telemedicine and eMedicine: a story from the RFID Center of Excellence at the University of Pittsburgh. Ann Biomed Eng. 2013;41(9):1913–1925.

15. Fife RS. Development of a comprehensive women’s health program in an academic medical center: experiences of the Indiana University National Center of Excellence in Women’s Health. J Womens Health (Larchmt). 2003;12(9):869–878.

16. JDRF News Column. New stem cell centre of excellence opens in Cambridge. Pract Diab Int. 2004; 21: 276–276. https://doi.org/10.1002/pdi.683.

17. Singh M, Sawarkar D, Sharma BS. Neurosurgery at All India Institute of Medical Sciences, a center of excellence: A success story. Neurol India. 2015;63(4):589–596.

18. Santos-Moreno P, Alvis-Zakzuk NJ, Villarreal-Peralta L, Carrasquilla-Sotomayor M, Paternina-Caicedo A, Alvis-Guzmán N. A comprehensive care program achieves high remission rates in rheumatoid arthritis in a middle-income setting. Experience of a Center of Excellence in Colombia. Rheumatol Int. 2018;38(3):499–505.

19. Yao Q, Zhou G. Role of a center of excellence program in improving the quality of peritoneal dialysis--a Chinese experience. Perit Dial Int. 2014;34 Suppl 2(Suppl 2):S59–S62.

20. Rábago CA, Clouser M, Dearth CL, et al. The Extremity Trauma and Amputation Center of Excellence: Overview of the Research and Surveillance Division. Mil Med. 2016;181(S4):3–12.

21. Arksey H, O’Malley L. Scoping studies: towards a methodological framework. Int J Soc Res Methodol. 2005;8(1):19–32.

22. Levac D, Colquhoun H, O’Brien KK. Scoping studies: advancing the methodology. Implement Sci. 2010;5:69.

23. Tricco AC, Lillie E, Zarin W, et al. PRISMA Extension for Scoping Reviews (PRISMA-ScR): Checklist and Explanation. Ann Intern Med. 2018;169(7):467–473.

24. Gao JJ, Pazdur R. FDA Oncology Center of Excellence During COVID-19-Working for Patients With Cancer. JAMA Oncol. 2020;10.1001/jamaoncol.2020.6783.

25. The University of Texas Health Science Center at Houston (UTHealth). COVID-19 Center of Excellence. Texas, USA; UTHealth 2020. https://www.uth.edu/covid-19/

26. Bialous SA, Nohavova I, Kralikova E, Wells MJ, Brook J, Sarna L. Building capacity in tobacco control by establishing the Eastern Europe Nurses’ Center of Excellence for Tobacco Control. Tob Prev Cessat. 2020;6:68.

27. Lipsitz O, McIntyre RS, Rodrigues NB, et al. Does body mass index predict response to intravenous ketamine treatment in adults with major depressive and bipolar disorder? Results from the Canadian Rapid Treatment Center of Excellence. CNS Spectr. 2020;1–9.

28. Komninaka V, Flevari P, Marinakis T, Karkaletsis G, Malakou L, Repa K. Outcomes of pregnancies in patients with Gaucher Disease: The experience of a center of excellence on rare metabolic Disease-Gaucher Disease, in Greece. Eur J Obstet Gynecol Reprod Biol. 2020;254:181–187.

29. Araujo-Castro M, Pascual-Corrales E, Martínez San Millan JS, et al. Postoperative management of patients with pituitary tumors submitted to pituitary surgery. Experience of a Spanish Pituitary Tumor Center of Excellence. Endocrine. 2020;69(1):5–17.

30. Balkhy HH, Amabile A, Torregrossa G. A Shifting Paradigm in Robotic Heart Surgery: From Single-Procedure Approach to Establishing a Robotic Heart Center of Excellence. Innovations (Phila). 2020;15(3):187–194.

31. Tinc PJ, Wolf-Gould C, Wolf-Gould C, Gadomski A. Longitudinal Use of the Consolidated Framework for Implementation Research to Evaluate the Creation of a Rural Center of Excellence in Transgender Health. Int J Environ Res Public Health. 2020;17(23):9047.

32. Sarkar T, Karmakar N, Dasgupta A, Saha B. Quality of life of people living with HIV/AIDS attending antiretroviral clinic in the center of excellence in HIV care in India. J Educ Health Promot. 2019;8:226.

33. Neal C, Rusangwa C, Borg R, et al. Cost of Treating Pediatric Cancer at the Butaro Cancer Center of Excellence in Rwanda. J Glob Oncol. 2018;4:1–7.

34. Choque-Velasquez J, Colasanti R, Baffigo-Torre V, et al. Developing the First Highly Specialized Neurosurgical Center of Excellence in Trujillo, Peru: Work in Progress-Results of the First Four Months. World Neurosurg. 2017;102:334–339.

35. Elrod JK, Fortenberry JL Jr. Centers of excellence in healthcare institutions: what they are and how to assemble them. BMC Health Serv Res. 2017;17(Suppl 1):425.

36. Santos-Moreno P, Caballero-Uribe CV, Massardo ML, et al. Systematic and progressive implementation of the centers of excellence for rheumatoid arthritis: a methodological proposal. Clin Rheumatol. 2017;36(12):2855–2858.

37. Chick JFB, Reddy SN, Pyeritz RE, Trerotola SO. A Survey of Pulmonary Arteriovenous Malformation Screening, Management, and Follow-Up in Hereditary Hemorrhagic Telangiectasia Centers of Excellence. Cardiovasc Intervent Radiol. 2017;40(7):1003–1009.

38. Perdomo HA. Cancer Centers of Excellence in Colombia: A fundamental way to work together. Urología colombiana. 2017;26(3):157–8.

39. Nwafor IA, Chinawa JM, Adiele DK, et al. Management of complex CHD at the National Cardiothoracic Center of Excellence, University of Nigeria Teaching Hospital, Enugu: the role of foreign cardiac missions in 3.5 years. Cardiol Young. 2017;27(6):1174–1179.

40. Casanueva FF, Barkan AL, Buchfelder M, et al. Criteria for the definition of Pituitary Tumor Centers of Excellence (PTCOE): A Pituitary Society Statement. 2018 Sep 20;:]. Pituitary. 2017;20(5):489–498.

41. Cohen MS. Enhancing surgical innovation through a specialized medical school pathway of excellence in innovation and entrepreneurship: Lessons learned and opportunities for the future. Surgery. 2017;162(5):989–993.

42. Doumouras AG, Saleh F, Anvari S, Gmora S, Anvari M, Hong D. A Longitudinal Analysis of Short-Term Costs and Outcomes in a Regionalized Center of Excellence Bariatric Care System. Obes Surg. 2017;27(11):2811–2817.

43. Scheuermann JS, Reddin JS, Opanowski A, et al. Qualification of National Cancer Institute-Designated Cancer Centers for Quantitative PET/CT Imaging in Clinical Trials. J Nucl Med. 2017;58(7):1065–1071.

44. Miranda JJ, Bernabé-Ortiz A, Diez-Canseco F, et al. Towards sustainable partnerships in global health: the case of the CRONICAS Centre of Excellence in Chronic Diseases in Peru. Global Health. 2016;12(1):29.

45. Kouchoukos NT. What is a Cardiothoracic Surgical “Center of Excellence”?. Ann Thorac Surg. 2016;102(5):1426–1427.

46. Melissas J, Stavroulakis K, Tzikoulis V, et al. Sleeve Gastrectomy vs Roux-en-Y Gastric Bypass. Data from IFSO-European Chapter Center of Excellence Program. Obes Surg. 2017;27(4):847–855.

47. Singh M, Sawarkar D, Sharma BS. Neurosurgery at All India Institute of Medical Sciences, a center of excellence: A success story. Neurol India. 2015;63(4):589–596.

48. Rábago CA, Clouser M, Dearth CL, et al. The Extremity Trauma and Amputation Center of Excellence: Overview of the Research and Surveillance Division. Mil Med. 2016;181(S4):3–12.

49. Rani PK, Balakrishnanan D, Padhi TR, Jalali S. Role of Retinopathy of Prematurity (ROP) Tertiary Centers of Excellence in Capacity-building. Indian Pediatr. 2016;53 Suppl 2:S85–S88.

50. Mettrick J, Harburger DS, Kanary PJ, et al. Building Cross-System Implementation Centers: A Roadmap for State and Local Child Serving Agencies in developing Centers of Excellence (COE). Baltimore, MD: The Institute for Innovation & Implementation, University of Maryland, 2015.

51. Hershko A, Edri MM, Wirtheim E. Harefuah. 2015;154(8):478–542.

52. Mehrotra A, Dimick JB. Ensuring excellence in centers of excellence programs. Ann Surg. 2015;261(2):237–239.

53. Katz IT, Bogart LM, Cloete C, et al. Understanding HIV-infected patients’ experiences with PEPFAR-associated transitions at a Centre of Excellence in KwaZulu Natal, South Africa: a qualitative study. AIDS Care. 2015;27(10):1298–1303.

54. Itri JN, Bakow E, Woods J. Creating an outpatient center of excellence in CT. J Am Coll Radiol. 2014;11(12 Pt A):1137–1143.

55. Reichert J, Furlong G. Five key pillars of an analytics center of excellence, which are required to manage populations and transform organizations into the next era of health care. Nurs Adm Q. 2014;38(2):159–165.

56. Zar HJ. Partnering with centers of excellence in high-and low-middle-income countries: a strategy to strengthen child health globally. Pediatr Radiol. 2014;44(6):709–710.

57. Ferguson GM, Froehlich JA. Establishing a center of excellence: the Total Joint Center at the Miriam Hospital. R I Med J (2013). 2013;96(3):16–18.

58. Mehrotra A, Sloss EM, Hussey PS, Adams JL, Lovejoy S, SooHoo NF. Evaluation of a center of excellence program for spine surgery. Med Care. 2013;51(8):748–757.

59. National Institutes of Health. NIH centers of excellence: Biennial Report of the Director, National Institutes of Health, Fiscal Years 2008 & 2009. publication no. 11-7701 volume 4. USA, NIH; 2009. https://report.nih.gov/biennialreport/

60. Sharkey K, Meeks-Sjostrom D, Baird M. Challenges in sustaining excellence over time. Nurs Adm Q. 2009;33(2):142–147.

61. Cullen L, Greiner J, Greiner J, Bombei C, Comried L. Excellence in evidence-based practice: organizational and unit exemplars. Crit Care Nurs Clin North Am. 2005;17(2):127–x.

62. Keto JL, Kemmeter PR. Effect of Center of Excellence requirement by Centers for Medicare and Medicaid Services on practice trends. Surg Obes Relat Dis. 2008;4(3):437–440.

63. Anderson RT, Weisman CS, Scholle SH, Henderson JT, Oldendick R, Camacho F. Evaluation of the quality of care in the clinical care centers of the National Centers of Excellence in Women’s Health. Womens Health Issues. 2002;12(6):309–326.

64. Pennsylvania Department of Human Services (DHS). Centers of excellence: Thinking about addiction a different way. Pennsylvania, USA; Pennsylvania DHS.

65. Dieye B, Affara M, Sangare L, et al. West Africa International Centers of Excellence for Malaria Research: Drug Resistance Patterns to Artemether-Lumefantrine in Senegal, Mali, and The Gambia. Am J Trop Med Hyg. 2016;95(5):1054–1060.

66. Nwaka S, Ochem A, Besson D, et al. Analysis of pan-African Centres of excellence in health innovation highlights opportunities and challenges for local innovation and financing in the continent. BMC Int Health Hum Rights. 2012;12:11.

67. Fekadu A, Hailu A, Makonnen E, Belete A, Yimer G. Short-term impact of celebrating the international clinical trial day: experience from Ethiopia. Trials. 2017;18(1):332.

68. McCarthy J. ESRF: A Quest for Excellence in Service to Users. Synchrotron Radiation News. 2017;30(1):31–6.

69. Heining C, von Kalle C. What functions do oncological centers of excellence have in future research?. ONKOLOGE. 2016 Mar 1;22(3):152–7.

70. Bloch C, Schneider JW, Sinkjær T. Size, Accumulation and Performance for Research Grants: Examining the Role of Size for Centres of Excellence. PLoS One. 2016;11(2):e0147726.

71. Borlaug SB. Moral hazard and adverse selection in research funding: Centres of excellence in Norway and Sweden. Science and Public Policy. 2016;43(3):352–62.

72. Pislyakov V, Shukshina E. Measuring excellence in Russia: Highly cited papers, leading institutions, patterns of national and international collaboration. Journal of the Association for Information Science and Technology. 2014;65(11):2321–30.

73. Househ M, Al-Tuwaijri M, Al-Dosari B. Establishing an Electronic Health Center of Research Excellence (E-CoRE) within the Kingdom of Saudi Arabia. Journal of Health Informatics in Developing Countries. 2010;4(1).

74. Lin CC, Yang CH, Cheng AL, Chan WK, Ho HN. National Center of Excellence for Clinical Trials and Research at National Taiwan University Hospital. Drug Information Journal. 2009;43(3):361–3.

75. Rocco G, Affonso D, Mayberry L, et al. Center of Excellence to build nursing scholarship and improve health care in Italy. Journal of Nursing Scholarship. 2015;47(2):170–7.

76. Fetrow J, Royster E, Morin D, et al. Development and Implementation of a National Center of Excellence in Dairy Production Medicine Education for Veterinary Students: Description of the Effort and Lessons Learned. J Vet Med Educ. 2020;47(3):250–262.

77. Harris TB, Salihu HM. 2019 Health Equity Summer Research Summit Organized by the Center of Excellence in Health Equity, Training and Research, Baylor College of Medicine, Houston, Texas 77030, USA on June 18th, 2019. Int J MCH AIDS. 2020;9(Suppl 1):S1-S47.

78. The World Bank. Eastern and Southern Africa Higher Education Centers of Excellence. Washington, D.C., USA; World Bank 2021. https://projects.worldbank.org/en/projects-operations/project-detail/P151847

79. Nwafor IA, Chinawa JM, Adiele DK, et al. Management of complex CHD at the National Cardiothoracic Center of Excellence, University of Nigeria Teaching Hospital, Enugu: the role of foreign cardiac missions in 3.5 years. Cardiol Young. 2017;27(6):1174–1179.

80. Kumar S, Abowd G, Abraham WT, et al. Center of Excellence for Mobile Sensor Data-to-Knowledge (MD2K). IEEE Pervasive Comput. 2017;16(2):18–22.

81. Giacomini KM, Lin L, Altman RB. Research Projects Supported by the University of California, San Francisco-Stanford Center of Excellence in Regulatory Science and Innovation. Clin Pharmacol Ther. 2019;105(4):815–818.

82. Terama E, Smallman M, Lock SJ, Johnson C, Austwick MZ. Beyond Academia - Interrogating Research Impact in the Research Excellence Framework. PLoS One. 2016;11(12):e0168533.

83. Cofrancesco J, Barone MA, Serwint JR, Goldstein M, Westman M, Lipsett PA. Development and Implementation of a School-Wide Institute for Excellence in Education to Enable Educational Scholarship by Medical School Faculty. Teach Learn Med. 2018;30(1):103–111.

84. Voznesenskaya A, Bougrov V, Kozlov S, Vasilev V. ITMO Photonics: center of excellence. InOptics Education and Outreach IV 2016 Sep 27 (Vol. 9946, p. 99460V). International Society for Optics and Photonics.

85. Bienenstock A, Schwaag Serger S, Benner M, Lidgard A. Combining excellence in education, research and impact: Inspiration from Stanford and Berkeley and implications for Swedish universities. SNS Forlag. 2014.

86. Rugen KW, Watts SA, Janson SL, et al. Veteran Affairs Centers of Excellence in Primary Care Education: transforming nurse practitioner education. Nurs Outlook. 2014;62(2):78–88.

87. Bittner NP, Ericson K, McCarthy M, Jones J. FOCUS on excellence: making a difference through global initiatives. Nurs Educ Perspect. 2013;34(1):25–28.

88. Balaji R, Venkadasalam S. Developing a marine engineering centre of excellence for competency-based training. WMU Journal of Maritime Affairs. 2017 May;16(2):293–311.

89. Sobhieh MH, Karimi R, Razmi J, et al. Develop Center of Excellence (COE) at Power Plant Projects in Southern Iran with Conceptual Model Approach to Supply Chain Management in Construction Industry Projects. Journal of Engineering and Applied Sciences. 2016;11:967–975.

90. Geiger J. Establishing a Center of Excellence. Information Management. 2006 Aug 1;16(8):24.

91. Nqampoyi V, Seymour LF, Laar DS. Effective business process management centres of excellence. InInternational Conference on Research and Practical Issues of Enterprise Information Systems 2016 (pp. 207–222). Springer, Cham.

92. Neely JR, de Supinski BR. Application modernization at LLNL and the Sierra Center of Excellence. Computing in Science & Engineering. 2017 Sep 1;19(5):9–18.

93. Wang M. CAS center for excellence in quantum information and quantum physics: Exploring frontiers of quantum physics and quantum technology. National Science Review. 2017 Jan 1;4(1):144–52.

94. Craig W, Fisher M, Garcia S, Kaylor C, Porter J, Reed LS. Generalized Criteria and Evaluation Method for Center of Excellence: A Preliminary Report. CARNEGIE-MELLON UNIV PITTSBURGH PA SOFTWARE ENGINEERING INST; 2009.

95. Grant LA, Gulsvig J, Call J. Measuring excellence: the new quality agenda. Provider. 2006;32(10):1–7.

96. Patel J, Andrews L. SEVEN KEY STEPS TO ESTABLISHING A CENTER OF EXCELLENCE-If you’re struggling to optimize your return on investment from Enterprise Content Management and other strategies, this just might offer a solution. Infonomics. 2010:40.

97. Marciniak R. Center of Excellence as a next step for shared service center. Journal of International Scientific Publication: Economy & Business, ISSN. 2012:1313-2555.

98. Hellström T. Centres of Excellence as a Tool for Capacity Building. Case Study: Canada. IMHE Higher Education Programme: Organization for Economic Cooperation and Development. 2013.

99. Brusoni M, Damian R, Sauri JG, et al. The concept of excellence in higher education. Retrieved on March. 2014;18:2016.

100. Heyes A. An Assessment of the Nuclear Security Centers of Excellence. Stanley Foundation; 2012.

101. Antonowicz D, Kohoutek J, Pinheiro R, Hladchenko M. The roads of ‘excellence in Central and Eastern Europe. European Educational Research Journal. 2017 Sep;16(5):547–67.

102. NYC Health + Hospitals. Mayor De Blasio and Taskforce on Racial Inclusion and Equity Announce New COVID-19 Centers of Excellence. New York, USA; NYC Health + Hospitals 2020. https://www.nychealthandhospitals.org/pressrelease/mayor-announces-three-new-covid-19-centers-of-excellence-gotham-health-sites/

103. Addis Ababa Science and Technology University (AASTU). The Artificial Intelligence & Robotics Center of Excellence. Addis Ababa, Ethiopia; AASTU 2021. http://www.aastu.edu.et/research-and-technology-transfer-vpresident/the-artificial-intelligence-robotics-center-of-excellence/

